# Cardiovascular safety and efficacy of combination therapy with metformin and sodium-glucose cotransporter-2 inhibitors versus metformin and sulfonylureas in patients with type 2 diabetes: a systematic review and meta-analysis of randomized controlled trials

**DOI:** 10.1101/2020.04.24.20077917

**Authors:** Desye Gebrie, Desalegn Getnet, Tsegahun Manyazewal

## Abstract

**Background:** Diabetes mellitus is a serious threat to global health and among the top 10 causes of death, with nearly half a billion people living with it worldwide. Treating patients with diabetes tend to become more challenging due to the nature of the disease. The role and benefits of combination therapies for the management of type 2 diabetes are well-documented, while the comparative safety and efficacy among the different combination options have not been elucidated. We aimed to systematically synthesize the evidence on the comparative cardiovascular safety and efficacy of combination therapy with metformin and sodium-glucose cotransporter-2 inhibitors versus metformin and sulfonylureas in patients with type 2 diabetes.

**Method:** We searched MEDLINE-PubMed, Embase, Cochrane Library, and ClinicalTrials.gov up to 15 August 2019 and without restriction in the year of publication. We included randomized controlled trials of patients with type 2 diabetes who were on metformin- sodium-glucose cotransporter-2 inhibitors or metformin-sulphonylureas combination therapy at least for a year. The primary endpoints were all-cause mortality and serious adverse events, and the secondary endpoints were cardiovascular mortality, non-fatal myocardial infarction, non-fatal stroke, hypoglycemia, and changes in glycosylated hemoglobin A1C, body weight, fasting plasma glucose, blood pressure, high-density lipoprotein cholesterol, and low-density lipoprotein cholesterol. Random effect model was carried out to calculate mean differences for continuous outcomes and risk ratio for dichotomous outcomes between the two combination therapies.

**Results:** Of 3,190 citations, we included nine trials involving 10,974 participants. The pooled analysis showed no significance difference in all-cause mortality (RR = 0.93, 95% CI [0.52, 1.67]), serious adverse events (RR=0.96, 95% CI [0.79, 1.17]) and adverse events (RR=1.00, 95% CI [0.99, 1.02]) between the two, but in hypoglycemia (RR= 0.13, 95% CI [0.10, 0.17], P<0.00001). Participants taking metformin-sodium-glucose cotransporter-2 inhibitors showed a significantly greater reduction in HbA1c (MD= −0.10, 95% CI [−0.17, −0.03] %, body weight (MD= −4.57, 95% CI [−4.74, −4.39] kg, systolic blood pressure (MD=-4.77, 95% CI [−5.39, −4.16] mmHg, p<0.00001), diastolic blood pressure (MD=-2.07, 95% CI [−2.74, −1.40] mmHg, and fasting plasma glucose (MD= −0.55, 95% CI [−0.69, −0.41] mmol/L, p < 0.00001).

**Conclusion:** A combination therapy of metformin and sodium-glucose cotransporter-2 inhibitors is a safe and efficacious alternative to combination therapy of metformin and sulphonylureas for patients with type 2 diabetes who are at risk of cardiovascular comorbidity. However, there remains a need for additional long-term randomized controlled trials as available studies are very limited and heterogeneous.

## Introduction

Diabetes mellitus (DM) is one of the top ten causes of death and one of the fastest growing health problems of the twenty-first century, with 463 million people living with it worldwide in 2019 and this number estimated to be 700 million by 2045 [1]. The global direct health expenditure on diabetes mellitus in 2019 was estimated US$ 760 billion and is expected to increase to a projected US$ 845 billion by 2045 [2]. Type 2 diabetes mellitus (T2DM) is the most common and complex form of the disease and accounts for more than 90% of the estimated cases of diabetes, impacting the life expectancy, quality of life, and health of an individual [1,3]. Yet, there is no cure for the T2DM, while its prevalence is largely increasing, with increased risk of complications including diabetic retinopathy, neuropathy, kidney damage, and microvascular and cardiovascular complications [4-7]. Cardiovascular disease (CVD) is a common complication and a major cause of morbidity and mortality in patients with T2DM [8,9].

Despite the introduction of new medications, treating patients with diabetes tend to become more challenging due to the nature of the disease [10-14]. The American Diabetes Association recommends lifestyle interventions (exercise, healthy eating, smoking cessation and weight reduction) as the first step in treating newly diagnosed patients with T2DM [15]. However, to achieve and maintain specific glycemic targets, the majority of patients require pharmacological glucose-lowering drugs. Metformin is currently the first-line and widely used pharmacological therapy for patients with T2DM because of its potential benefits, including cardioprotective effect, loss of weight and prevention of some comorbid diseases [15-22]. If lifestyle interventions and a maximal tolerated dose of metformin fail to achieve the glycemic target within 3 months follow-up, the regimen would be changed to combination therapy [15].

Metformin-sulfonylurea combination therapy is the most widely used regimen in T2DM [23, 24]. Sulfonylureas are prescribed as second-line treatment options in the management of patients with T2DM, while they are still commonly prescribed also as a first-line treatment instead of metformin [25]. However, initiating treatment of T2DM with a sulfonylurea rather than metformin is associated with higher rates of ischaemic stroke, cardiovascular mortality, and hypoglycemia [25-31]. Besides, sulfonylureas as second-line treatments are associated with an increased risk of non-fatal myocardial infarction, all-cause mortality, and severe hypoglycemia, compared with metformin monotherapy; as a result, continuing metformin when introducing sulfonylureas appears to be safer than switching [32]. Such findings led to new requirements from licensing authorities that all new T2DM therapies should show cardiovascular safety [10].

Sodium-glucose cotransporter-2 inhibitors (SGLT2Is) are novel antidiabetic drugs that can inhibit sodium-glucose cotransporter-2 at the proximal tubule of the kidney. Those novel drugs can decreases renal glucose reabsorption, hyperglycemia, cardiovascular problems and they can increases urinary glucose excretion in patients with T2DM [33-39]. However, there is no clear evidence which shows the relative advantage of metformin with sulphonylureas or metformin with another glucose-lowering drugs on major treatment outcomes including CVD [40]. Management in T2DM has become challenging. Because, choosing a second and/or third-line antidiabetic drug is personalized based on efficacy, risk of hypoglycemia, patient’s comorbid conditions, impact on weight, side effects, and cost [41]. In particular, although most patients with T2DM require a combination pharmacological therapy, the choice of a best second-line drug is especially critical for the prevention of CVD.

Thus, the aim of this systematic review and meta-analysis of randomized controlled trials (RCTs) was to compare the cardiovascular safety and efficacy of a combination therapy of metformin-SGLT2I and metformin-sulfonylureas in patients with T2DM.

## Methods

The protocol for this systematic review and meta-analysis was submitted for registration in the PROSPERO database of prospectively registered systematic reviews and is awaiting the registration. We followed the Preferred Reporting Items for Systematic Review and Meta-Analysis (PRISMA 2015) guidelines [42] for the design and reporting of the results.

### Data sources and searches

We searched MEDLINE/PubMed (http://www.ncbi.nlm.nih.gov/pubmed/), Embase (http://www.embase.com/), The Cochrane Library (http://www.cochranelibrary.com/), ClinicaTtrials.gov (https://www.clinicaltrials.gov/), and google scholar (https://scholar.google.com/) databases for completed studies that reported the safety and/or efficacy of metformin-SGLT2I versus metformin-sulfonylureas combination therapies for patients with T2DM. We included RCTs without restriction in year of publication, but published in English language, up to 15 August 2019. The RCTs were needed to have at least a one-year follow-up of patients. The Medical Subject Headings (MeSH) and keywords we used in different combinations using balloon operators were: metformin, biguanide, sodium-glucose co-transporter-2 inhibitor, SGLT-2 inhibitor, dapagliflozin, canagliflozin, empagliflozin, ertugliflozin, sulfonylurea, gliclazide, glimepiride, glyburide, glibenclamide, glipizide, tolbutamide, type 2 diabetes mellitus, T2DM, cardiovascular outcomes, and randomized controlled trials. All potentially eligible studies were considered for this review, irrespective of the primary outcomes. Manual searching was performed to find out additional eligible trials from the reference lists of key articles.

### Eligibility

The study eligibility criteria for this systematic review and meta-analysis were in accordance with the PICOS (participants, interventions, comparison, outcomes, and study designs) models[43].

#### Participants

The participants were patients with T2DM who were taking a combination therapy of metformin and sulfonylurea or metformin and SGLT2Is.

#### Intervention

Intervention was a combination of metformin and any of the SGLT2I, which could be dapagliflozin, canagliflozin, empagliflozin, or ertugliflozin.

#### Comparator

Comparator was a combination of metformin and any of sulfonylureas compounds, which could be gliclazide, glipizide, glyburide, glibenclamide, or glimepiride.

#### Outcomes

The primary endpoints were all-cause mortality and serious adverse events (SAEs). The secondary endpoints were cardiovascular mortality, non-fatal myocardial infarction, non-fatal stroke, hypoglycemia, and changes in glycosylated hemoglobin A1C (HbA1c), body weight, fasting plasma glucose (FPG), systolic blood pressure (SBP), diastolic blood pressure (DBP), low-density lipoprotein cholesterol (LDL-C), and high-density lipoprotein cholesterol (HDL-C).

#### Study design

Only RCTs were needed to have at least a one-year follow-up of patients.

### Study selection

The title and abstract of all searched studies were examined by two independent review authors DG (first author) and DG (second author). From the title and abstract of all studies identified by the database search, those studies duplicated and not meet the inclusion criteria were excluded. The full texts of the remaining studies were further reviewed. Disagreements were resolved by consensus and if persisted, we were arbitrated through discussion with a third review author (TM).

### Data extraction

The following data were extracted from each included RCTs: first author, year of publication, mean age of the participant, baseline average body weight, HbA1c level of the participant, interventions, comparators, number of participants randomized, duration of follow-up, and patient-important outcomes. Data on the mean change of HbA1c (%), body weight (Kg), FPG (mmol/L), SBP (mmHg), DBP (mmHg), HDL-C (mmol/L) and LDL-C (mmol/L) were collected from baseline for continuous outcomes. The number of events (death, hypoglycemia, adverse events (AEs), AEs related to study drug, serious adverse events (SAEs), SAEs related to study drug, genital mycotic infection (GMI), cardiovascular events) were identified between the two combination arms.

### Assessment of risk of bias

The Cochrane risk of bias tool [44]was used to assess the risk of bias for each included study. The risk of bias of each trial was judged by two independent review authors as “Low”, “Unclear”, or “High” based on the critical domains, including random sequence generation, allocation concealment, blinding, incomplete outcome data, and selective reporting.

### Statistical analysis

Meta-analyses was carried out using the computer software packages RevMan 5.3 [45]. The meta-analysis compared the cardiovascular safety and efficacy of SGLT2I and sulfonylureas when combined with metformin as a second-line drug. Pooled results of continuous patient-important outcomes i.e., HbA1c, FPG, blood pressure, body weight, HDL-C, and LDL-C were reported using a mean difference (MD) with 95% confidence interval (CI). Pooled results of binary outcomes i.e. all-cause mortality, AEs, AEs related to study drug, SAEs, SAEs related to study drug, hypoglycemic event, worsening of coronary artery disease, acute myocardial infarction (AMI), aortic aneurism, coronary artery occlusion (CAO) and GMI were summarized using risk ratio (RR) with 95% CI. Mantel-Haenszel method [46] was used to pool effect estimates of dichotomous outcomes and inverse variance for continuous outcomes. The analysis was conducted using a random-effects model as it is preferable for smaller studies compared to a fixed-effect model meta-analysis [47]. Cochrane Q test [48] was used to assess heterogeneity between studies, and I^2^ testing [49] was done to quantify heterogeneity between studies, with values > 50% representing moderate-to-high heterogeneity. Sensitivity and subgroup analysis were carried out by the length of the trial duration. Statistical analysis with a p-value < 0.05 was considered statistically significant.

## Results

### Search results

A total of 3,190 citations were searched through the electronic databases, of which 30 full-text studies were assessed for eligibility and nine of these [50-58] fulfilled the inclusion criteria. We excluded the rest 21 full-text articles [23,24, 59-77] mainly for they did not include SGLT2I or sulfonylureas in the combination therapy; included a combination of more than two glucose-lowering drugs; included single glucose-lowering drug; they were driven from review or post hoc analysis of previous RCTs; they had no active comparator; or duration of interventions were less than a year (Figure 1).

**Figure 1:**
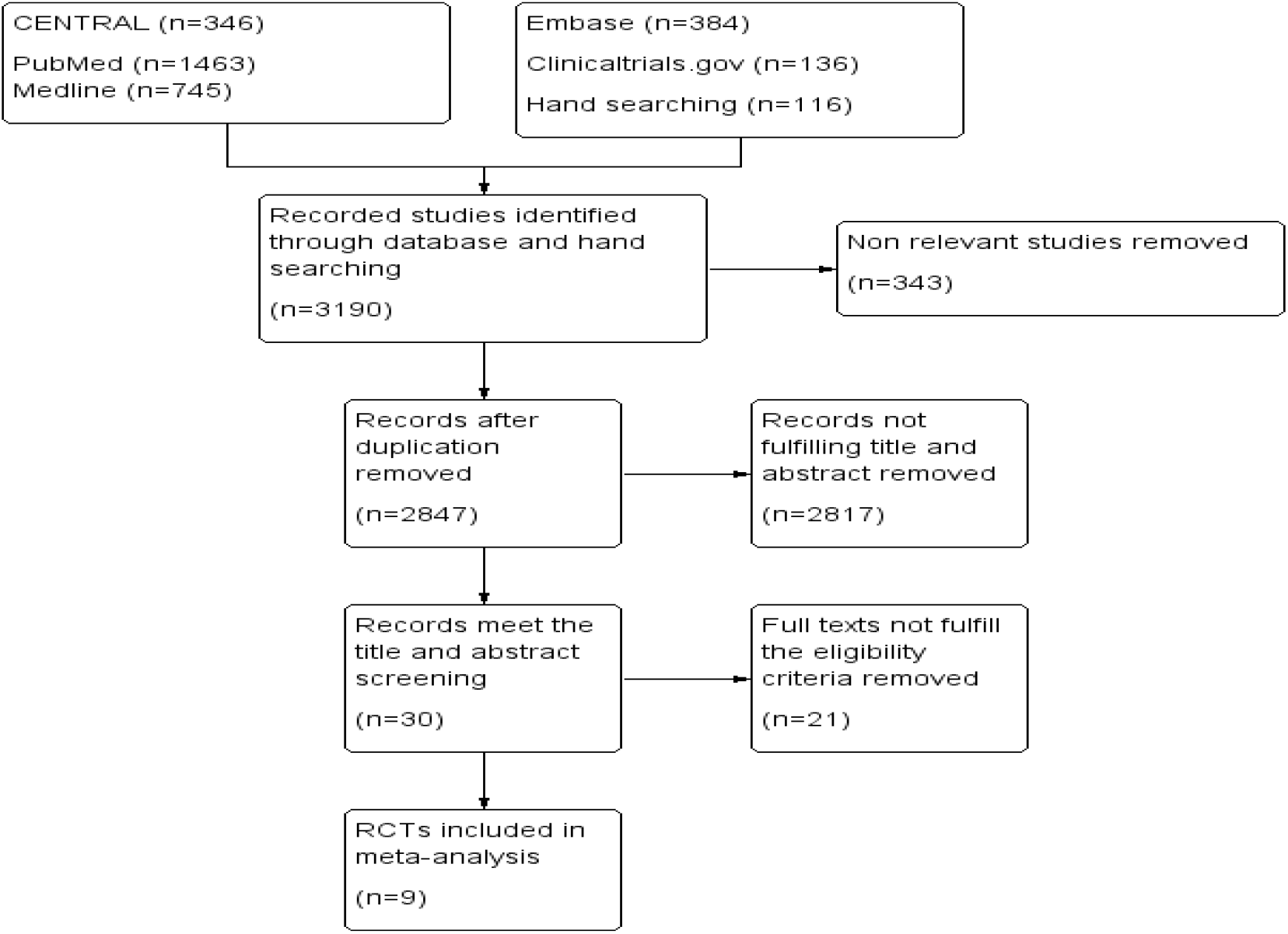
PRISMA flow diagram of the study selection process and search results

### Study characteristics

Table 1 summarizes the characteristics of the nine RCTs included. Four of the nine RCTs [50, 53, 55, 58] used two different doses of SGLT2I. The meta-analysis included all results of both doses for dichotomous outcomes but only a high dose of SGLT2I for continuous outcomes.

**Table 1:** characteristics of included RCTs

| 1<sup>st</sup>author (year) | Age (year) | Weight (Kg) | HbA1c (%) | No. of pts | Intervention | Comparator | Follow – up (year) | Patient important outcomes | Met+ SGLT2I | Met+SU |
| --- | --- | --- | --- | --- | --- | --- | --- | --- | --- | --- |
| Ridderstråle (2018) [51] | ≥18 | 82.75 | 7-10 | 1545 | Metformin plus Empagliflozin (n=765) | Metformin plus Glimepiride (n=780) | 4 | HbA1c (%)<br>weight (Kg)<br>SBP (mmHg)<br>DBP (mmHg)<br>FPG (mmol//L)<br>AEs<br>AEs related to drug<br>SAEs<br>SAEs related to drug<br>Hypoglycemia<br>Pt received rescue Rx<br>Dyslipidemia<br>GMI<br>CV events<br>Death | -0.29<br>-3.08<br>-4.1<br>-1.8<br>-0.3<br>706<br>221<br>161<br>N/A<br>23<br>176<br>↓fat<br>104<br>↔<br>8 | -0.10<br>1.84<br>2.1<br>0.8<br>0.4<br>713<br>277<br>153<br>N/A<br>218<br>265<br>↑fat<br>30<br>↑<br>8 |
| Ridderstråle (2014) [52] | ≥18 | 82.75 | 7-10 | 1545 | Metformin plus Empagliflozin (n=765) | Metformin plus Glimepiride (n=780) | 2 | HbA1c<br>FPG<br>Weight<br>SBP<br>DBP<br>AEs<br>AEs related to drug<br>SAEs<br>Hypoglycemia<br>Dyslipidemia<br>GMI<br>Pt's HbA1c<7%<br>Pt received rescue Rx<br>Death | -0.66<br>-0.85<br>-3<br>-3.1<br>-1.8<br>661<br>190<br>119<br>19<br>41<br>131<br>232<br>113<br>5 | -0.55<br>-0.17<br>1.5<br>2.5<br>0.9<br>673<br>252<br>89<br>189<br>39<br>22<br>221<br>185<br>5 |
| Hollander (2019) [53] | 58.2±9.6 | 86.9±19.6 | 7.8±0.6 | 1037 | Metformin ≥1500mg plus Ertugliflozin 15mg and | Metformin ≥1500mg plus Glimepiride 1-8mg (n=349) | 2 | HbA1c<br>Weight<br>FPG<br>SBP<br>DBP | -0.7<br>-6.3<br>-1.4<br>-3.2<br>-1.5 | -0.4<br>1.0<br>-0.5<br>2.1<br>0.7 |
|  |  |  |  |  | Metformin plus<br>Ertugliflozin 5mg<br>(n=688) |  |  | HDL-C | ↑ | ↓ |
|  |  |  |  |  |  |  |  | LDL-C | ↔ | ↔ |
|  |  |  |  |  |  |  |  | Hypoglycemia | 35 | 77 |
|  |  |  |  |  |  |  |  | AEs | 622 | 303 |
|  |  |  |  |  |  |  |  | SAEs | 73 | 30 |
|  |  |  |  |  |  |  |  | GMI | 67 | 3 |
|  |  |  |  |  |  |  |  | SAEs related to drug | 4 | 1 |
|  |  |  |  |  |  |  |  | Pt received rescue Rx | 60 | 31 |
|  |  |  |  |  |  |  |  | Pt's HbA1c <7% | 228 | 123 |
|  |  |  |  |  |  |  |  | Death | 9 | 1 |
| Hollander (2017)<br>[58] | 58.2±9.6 | 86.8±19.6 | 7.8±0.6 | 1326 | Metformin<br>≥1500mg plus<br>Ertugliflozin<br>15mg<br>And<br>Metformin plus<br>Ertugliflozin 5mg<br>(n=888) | Metformin ≥<br>1500mg plus<br>Glimepiride 1-8mg<br>(n=437) | 1 | HbA1c | -1.2 | -0.7 |
|  |  |  |  |  |  |  |  | FPG | -2.3 | -0.9 |
|  |  |  |  |  |  |  |  | SBP | -6.0 | 1.0 |
|  |  |  |  |  |  |  |  | DBP | -2.1 | 0.3 |
|  |  |  |  |  |  |  |  | Weight | -6.4 | 0.9 |
|  |  |  |  |  |  |  |  | Sever hypoglycemia | 2 | 10 |
|  |  |  |  |  |  |  |  | AEs | 525 | 269 |
|  |  |  |  |  |  |  |  | SAEs | 45 | 12 |
|  |  |  |  |  |  |  |  | Pt received rescue Rx | 41 | 14 |
|  |  |  |  |  |  |  |  | SAE related to drug | 3 | 1 |
|  |  |  |  |  |  |  |  | GMI | 56 | 3 |
|  |  |  |  |  |  |  |  | HDL | ↑ | ↓ |
|  |  |  |  |  |  |  |  | LDL | ↑ | ↓ |
|  |  |  |  |  |  |  |  | Atherosclerosis | 0 | 1 |
|  |  |  |  |  |  |  |  | History of PVD | 1 | 0 |
|  |  |  |  |  |  |  |  | Pt's HbA1c<7% | 321 | 190 |
|  |  |  |  |  |  |  |  | Death | 6 | 1 |
| Del Prato (2015)<br>[57] | 58.2 | 89.7 | 7.5 | 814 | Metformin plus<br>Dapagliflozin<br>(n=406) | Metformin<br>plus Glipizide<br>(n=408) | 4 | HbA1c (%) | -0.10 | 0.20 |
|  |  |  |  |  |  |  |  | FPG (mg/dl) | -13.3 | -3.8 |
|  |  |  |  |  |  |  |  | Weight (Kg) | -3.65 | 0.73 |
|  |  |  |  |  |  |  |  | SBP | -3.69 | -0.02 |
|  |  |  |  |  |  |  |  | DBP | N/A | N/A |
|  |  |  |  |  |  |  |  | HDL-C | N/A | N/A |
|  |  |  |  |  |  |  |  | LDL-C | N/A | N/A |
|  |  |  |  |  |  |  |  | AEs | 356 | 355 |
|  |  |  |  |  |  |  |  | AEs related to drug | 127 | 127 |
|  |  |  |  |  |  |  |  | SAEs | 75 | 81 |
|  |  |  |  |  |  |  |  | SAEs related to drug | 9 | 7 |
|  |  |  |  |  |  |  |  | Hypoglycemia | 4 | 46 |
|  |  |  |  |  |  |  |  | Poor glycemic control | 156 | 182 |
|  |  |  |  |  |  |  |  | Cardiovascular events | 0 | 0 |
|  |  |  |  |  |  |  |  | GMI | 5 | 3 |
|  |  |  |  |  |  |  |  | Death | 2 | 5 |
| Nauck (2014)<br>[56] | 58.4 | 88 | 7.7 | 814 | Metformin plus<br>Dapagliflozin<br>(n=406) | Metformin<br>plus Glipizide<br>(n=408) | 2 | HbA1c | -0.32 | -0.14 |
|  |  |  |  |  |  |  |  | FPG | -1.12 | -0.68 |
|  |  |  |  |  |  |  |  | Weight | -3.7 | 1.4 |
|  |  |  |  |  |  |  |  | SPB | -2.7 | 1.2 |
|  |  |  |  |  |  |  |  | DBP | N/A | N/A |
|  |  |  |  |  |  |  |  | HDL-C | N/A | N/A |
|  |  |  |  |  |  |  |  | LDL-C | N/A | N/A |
|  |  |  |  |  |  |  |  | AEs | 337 | 338 |
|  |  |  |  |  |  |  |  | AEs related to drug | 122 | 118 |
|  |  |  |  |  |  |  |  | SAEs | 51 | 62 |
|  |  |  |  |  |  |  |  | SAEs related to drug | 8 | 7 |
|  |  |  |  |  |  |  |  | Hypoglycemia | 17 | 187 |
|  |  |  |  |  |  |  |  | Aortic aneurysm | 0 | 1 |
|  |  |  |  |  |  |  |  | AMI | 1 | 1 |
|  |  |  |  |  |  |  |  | CAO | 0 | 1 |
|  |  |  |  |  |  |  |  | Ventricular arrhythmia | 1 | 0 |
|  |  |  |  |  |  |  |  | Worsening CAD | 1 | 0 |
|  |  |  |  |  |  |  |  | GMI | 60 | 12 |
|  |  |  |  |  |  |  |  | Poor glycemic control | 55 | 74 |
|  |  |  |  |  |  |  |  | Death | 0 | 4 |
| Nauck (2011)<br>[54] | 58.4 | 88 | 7.7 | 814 | Metformin plus<br>Dapagliflozin<br>(n=406) | Metformin<br>plus Glipizide<br>(n=408) | 1 | HbA1c (%) | -0.52 | -0.52 |
|  |  |  |  |  |  |  |  | FPG (mmol/L) | -1.12 | -1.59 |
|  |  |  |  |  |  |  |  | Weight (Kg) | -3.2 | 1.2 |
|  |  |  |  |  |  |  |  | SBP (mmHg) | -3.8 | 0.9 |
|  |  |  |  |  |  |  |  | DBP (mmHg) | ↓ | ↑ |
|  |  |  |  |  |  |  |  | HDL-C | ↑ | ↓ |
|  |  |  |  |  |  |  |  | AEs | 318 | 318 |
|  |  |  |  |  |  |  |  | AEs related to drug | 110 | 110 |
|  |  |  |  |  |  |  |  | SAEs | 35 | 46 |
|  |  |  |  |  |  |  |  | SAEs related to drug | 6 | 4 |
|  |  |  |  |  |  |  |  | Hypoglycemia | 14 | 162 |
|  |  |  |  |  |  |  |  | GMI | 50 | 11 |
|  |  |  |  |  |  |  |  | Poor glycemic control | 1 | 15 |
|  |  |  |  |  |  |  |  | Acute MI | 0 | 1 |
|  |  |  |  |  |  |  |  | Worsening CAD | 1 | 0 |
|  |  |  |  |  |  |  |  | Death | 0 | 3 |
| Leiter (2015)<br>[55] | 56.2 | 86.6 | 7.8 | 1450 | Metformin plus<br>Canagliflozin<br>100mg and<br>Metformin plus<br>canaglifloxin<br>300mg<br>(n=968) | Metformin<br>plus<br>Glimepiride<br>(n=482) | 2 | HbA1c (%) | -1.39 | -0.55 |
|  |  |  |  |  |  |  |  | FPG (mmol/L) | -2.4 | -0.6 |
|  |  |  |  |  |  |  |  | Weight (Kg) | -7.2 | 0.8 |
|  |  |  |  |  |  |  |  | SBP | -5.1 | 1.7 |
|  |  |  |  |  |  |  |  | DBP | -3.5 | -0.02 |
|  |  |  |  |  |  |  |  | LDL-C (mmol/L) | 0.38 | 0.06 |
|  |  |  |  |  |  |  |  | HDL-C | 0.21 | 0.00 |
|  |  |  |  |  |  |  |  | Triglycerides | ↑ | ↔ |
|  |  |  |  |  |  |  |  | Pt's HbA1c<7% | 448 | 212 |
|  |  |  |  |  |  |  |  | AEs | 732 | 378 |
|  |  |  |  |  |  |  |  | AEs related to drug | 297 | 134 |
|  |  |  |  |  |  |  |  | SAEs | 94 | 69 |
|  |  |  |  |  |  |  |  | Hypoglycemia | 73 | 197 |
|  |  |  |  |  |  |  |  | GMI | 116 | 11 |
|  |  |  |  |  |  |  |  | Death | 6 | 2 |
| Cefalu (2013)<br>[50] | 56.2 | 86.6 | 7.8 | 1450 | Metformin plus<br>Canagliflozin<br>100mg and<br>Metformin plus<br>canaglifloxin<br>300mg<br>(n=968) | Metformin<br>plus<br>Glimepiride<br>(n=482) | 1 | HbA1c | -1.75 | -0.81 |
|  |  |  |  |  |  |  |  | FPG | -2.87 | -1.02 |
|  |  |  |  |  |  |  |  | Weight | -7.7 | 0.7 |
|  |  |  |  |  |  |  |  | SBP | -7.9 | 0.2 |
|  |  |  |  |  |  |  |  | DBP | -4.3 | -0.1 |
|  |  |  |  |  |  |  |  | LDL-C | 0.37 | 0.05 |
|  |  |  |  |  |  |  |  | HDL-C | 0.18 | -0.01 |
|  |  |  |  |  |  |  |  | Triglycerides | -0.32 | -0.01 |
|  |  |  |  |  |  |  |  | Pt's HbA1c<7% | 541 | 264 |
|  |  |  |  |  |  |  |  | AEs | 643 | 330 |
|  |  |  |  |  |  |  |  | AEs related to drug | 263 | 113 |
|  |  |  |  |  |  |  |  | SAEs | 50 | 39 |
|  |  |  |  |  |  |  |  | Hypoglycemia | 51 | 165 |
|  |  |  |  |  |  |  |  | GMI | 97 | 8 |
|  |  |  |  |  |  |  |  | Death | 2 | 2 |

AEs=Adverse Events, AMI=Acute Myocardial Infraction, CAD=Coronary Artery Disease, CAO=Coronary Artery Occlusion, CV=Cardiovascular, HbA1c=Hemoglobin A1c, FPG= Fasting plasma
glucose, SBP= Systolic blood pressure, DBP=Diastolic blood pressure, HDL-C=High Density Lipoprotein Cholesterol, LDL-C=Low Density Lipoprotein Cholesterol, SAEs=Serious Adverse Events,
GMI=Genital Mycotic Infection, MI=Myocardial Infraction, Rx=Treatment, SGLT2I=Sodium Glucose Co-transporter 2 Inhibitor, SU=Sulfonylurea, Met=Metformin, N/A=Not Available,

### Participant characteristics

From the nine RCTs included, we polled and included 10,974 patients with type 2 diabetes who were on metformin-SGIT2I or metformin-sulphonylureas combination therapy at least for a year (Table 1).

### Methodological quality and risk of bias

The studies were found to be “low risk of bias” when these studies were subjected to the Cochrane Collaboration’s Tool for Quality Assessment of Randomized Controlled Trials (Figure 2).

**Figure 2:**
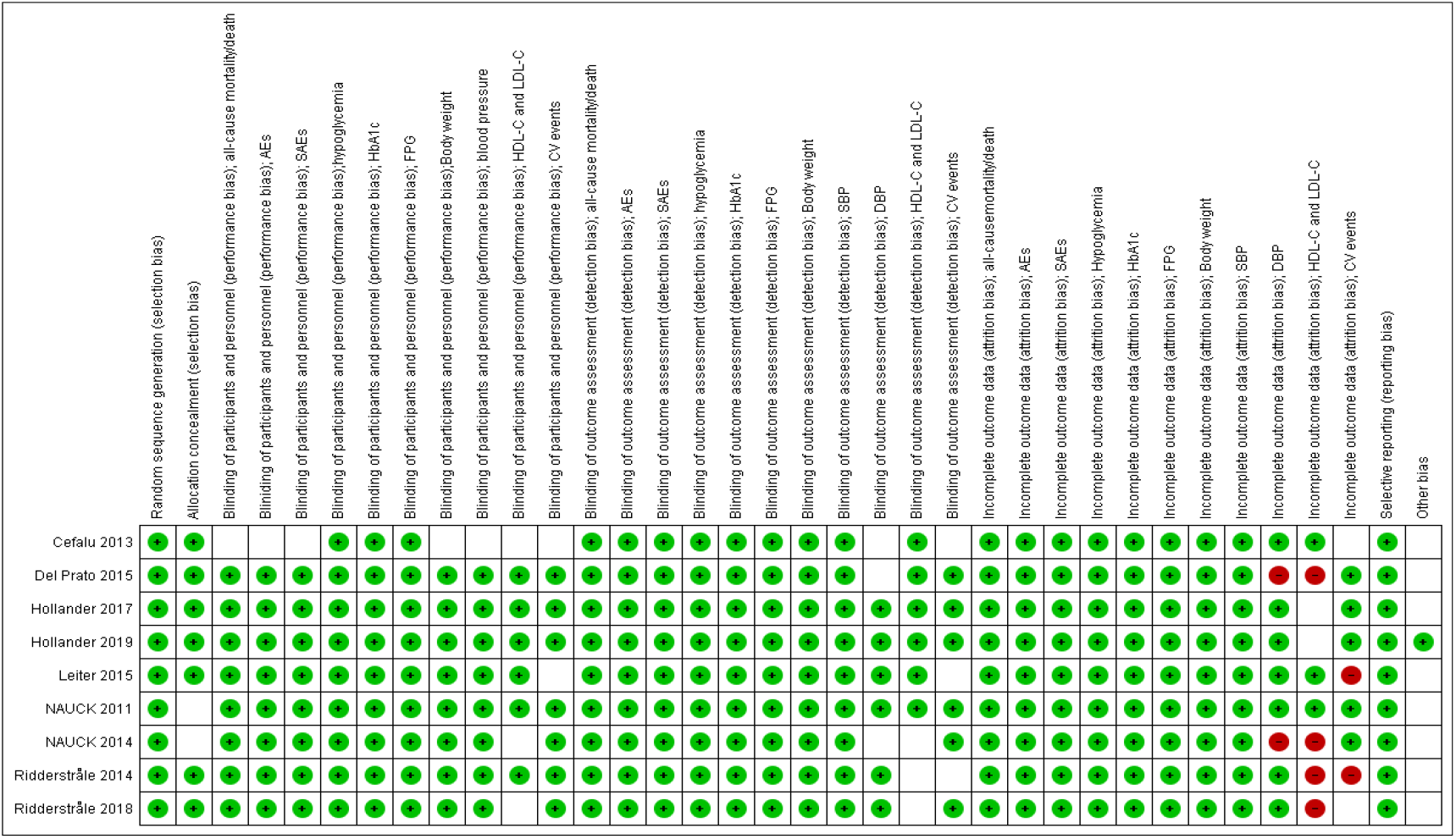
‘Risk of bias’ summary: review authors’ judgments about each ‘risk of bias’ item for included trials (Green cells = ‘low risk’; blank cells = ‘unclear risk’; red cells = ‘high risk’)

### Efficacy and safety assessments

#### All-cause mortality

All the nine included RCTs involving 10,974 participants assessed all-cause mortality/death events between intervention and control groups. Our meta-analysis of pooled results revealed no significance difference in all-cause mortality/death events between patients with T2DM who were on metformin-SGIT2I and metformin-sulphonylureas combination therapies (RR = 0.93, 95% CI [0.52, 1.67], *p* = 0.81), with statistically non-significant heterogeneity between studies (I^2^ = 9%) (Figure 3). Subgroup analysis by duration of follow-up showed no significant difference in risk of death between the two groups, both at follow-up year 1 (RR= 0.66, 95% CI [0.12, 3.71], p= 0.64, year 2 (RR=1.25, 95% CI [0.47,3.32], p= 0.66, and year 4 (RR=0.80, 95% CI [0.35, 1.84], p=0.60) (Appendix I).

**Figure 3:**
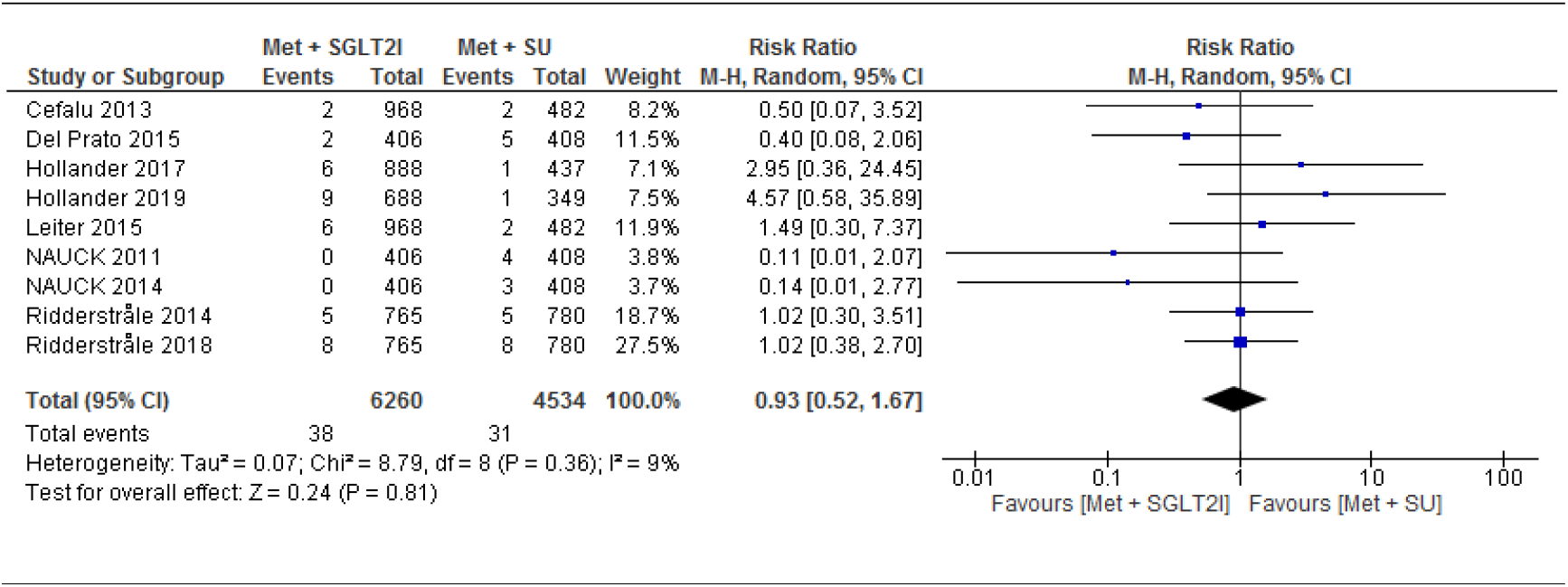
Comparison of all-cause mortality/death events between patients with T2DM who were on metformin-SGIT2I and metformin-sulphonylureas combination therapies. RR= Risk Ratio; CI= confidence interval; df= degrees of freedom; Met=Metformin; SGLT2I= Sodium-glucose co-transporter 2 inhibitor; SU= Sulfonylurea

#### Cardiovascular events

Two trials [54, 56] evaluated the cardiovascular efficacy of metformin-SGIT2I versus metformin-sulphonylureas combination therapies. The studies followed-up 1,628 participants for coronary artery disease (CAD) and acute myocardial infarction (AMI). After 2 years follow-up, there was no statistically significant difference observed for CAD (RR= 3.01, 95% CI [0.31, 28.92], p=0.34) and AMI (RR=0.63, 95% CI [0.08, 5.09], p=0.66) between the two arms. One of the two trials [56] further evaluated 814 patients for coronary artery occlusion (CAO) and aortic aneurism and reported no statistically significant difference in the risk of developing theses diseases between the two arms (Appendix II).

Of the nine trials included, eight trials followed-up 8,399 participants for changes in systolic blood pressure from baseline and five trials followed-up 5,804 participants for changes in diastolic blood pressure. The report showed a significant decreases for both systolic blood pressure (MD=-4.77, 95% CI [−5.39, −4.16] mmHg, p<0.00001) and diastolic blood pressure (MD=-2.07, 95% CI [−2.74, −1.40] mmHg, p<0.00001) in patients taking metformin-SGIT2I combination therapy (Figure 4 and 5).

**Figure 4:**
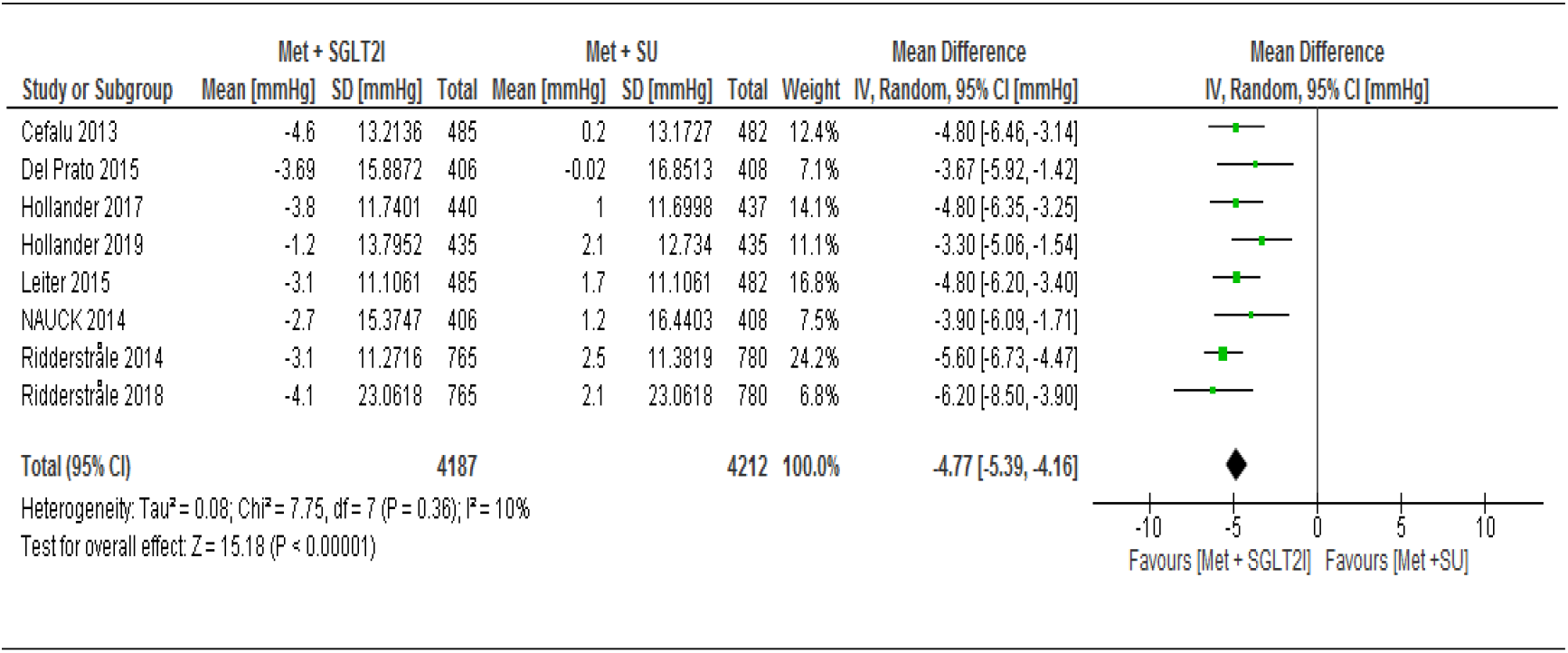
Comparison of changes in systolic blood pressure between patients who were on metformin-SGIT2I and metformin-sulphonylureas combination therapies. CI= confidence interval; df= Degree of freedom; MD= Mean difference; Met= Metformin; SGLT2I= Sodium-glucose co-transporter 2 inhibitor; SU= Sulfonylurea; SD= standard deviation

**Figure 5:**
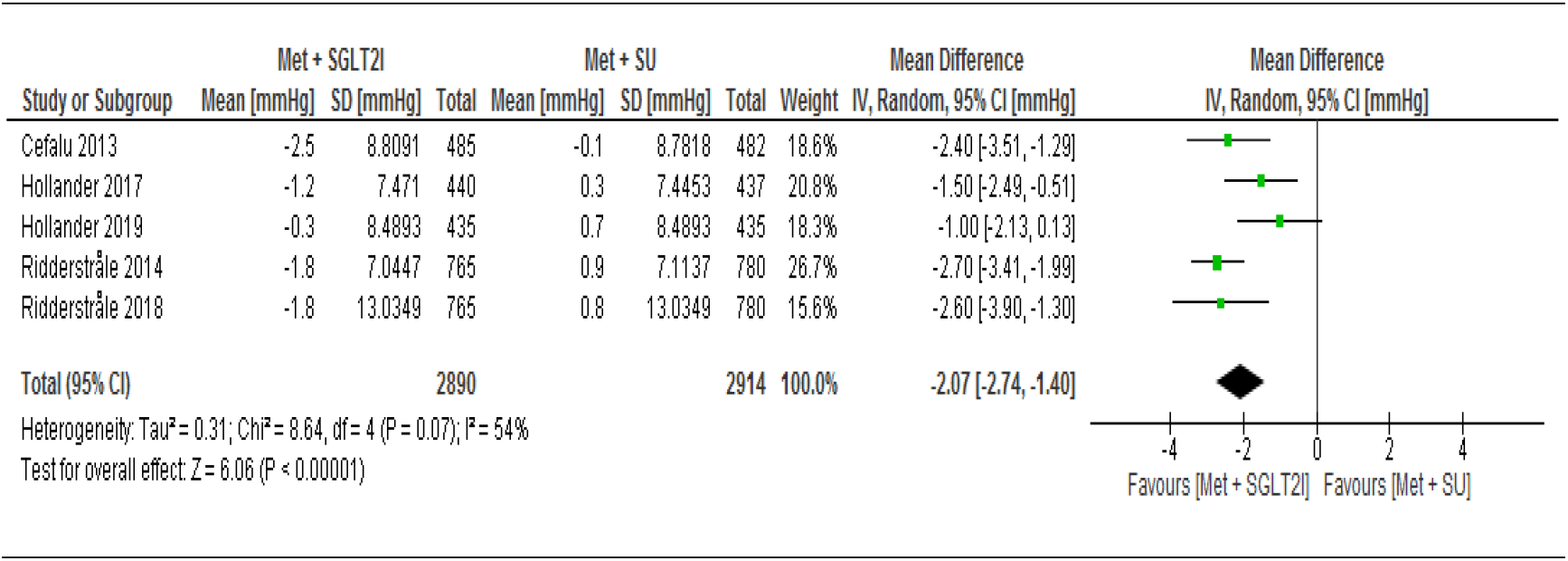
Comparison of changes in diastolic blood pressure between patients who were on metformin-SGIT2I and metformin-sulphonylureas combination therapies. CI= confidence interval; df= Degree of freedom; MD= Mean difference; Met=Metformin; SGLT2I= Sodium-glucose co-transporter 2 inhibitor; SU= Sulfonylurea; SD= standard deviation

Two trials [50, 55] assessed the change in HDL-C and LDL-C levels from baseline between the two arms. Both HDL-C and LDL-C levels reduced on patients on metformin-sulphonylureas combination therapy. However, the pooled effect was not statistically significant between the two arms for HDL-C (MD= 4.32, 95% CI [−4.00, 12.64] mmol/L, p=0.32), I^2^ =99%) and LDL-C (MD=3.63, 95% CI [−3.96, 11.22] mmol/L, P=0.35, I^2^= 88%).

#### Adverse events

With pooled data from the nine trials, we found no statistically significant difference between the two arms in the risk of developing adverse events (RR=1.00, 95% CI [0.99, 1.02], p=0.66, I^2^=0% (Figure 6). We performed a sensitivity analysis by removing the highest weighted study [51] and found no significant difference in the risk of developing adverse events between the two arms (RR=1.00, 95% CI [0.98, 1.02], I^2^ =0%). The result of the subgroup analysis was consistent at different duration of follow-up (RR=0.98, 95% CI [0.94, 1.03] at 1 year, RR=1.00, 95% CI [0.97, 1.04] at 2 years and RR=1.01, 95% CI [0.98, 1.04] at 4 years, with p=0.57 for subgroup difference (Appendix III). Seven RCTs assessed the risk of adverse events related to study drug, of which two RCTs showed lower risk in patients on metformin-sulphonylureas combination therapies but the pooled analysis showed no statistically significant difference (RR=0.97, 95% CI [0.85, 1.10], p=0.60, I^2^=69%, (Appendix IV). Subgroup analysis of the seven RCTs showed that risk of adverse events related to study drug was similar across different duration of study, with RR =1.09, 95% CI [0.94, 1.26], I^2^ = 0% at year one, RR = 0.94, 95% CI [0.75, 1.21], I^2^ = 81% at year two, and RR = 0.89, 95% CI [0.73, 1.10], I^2^ = 63% at year four.

**Figure 6:**
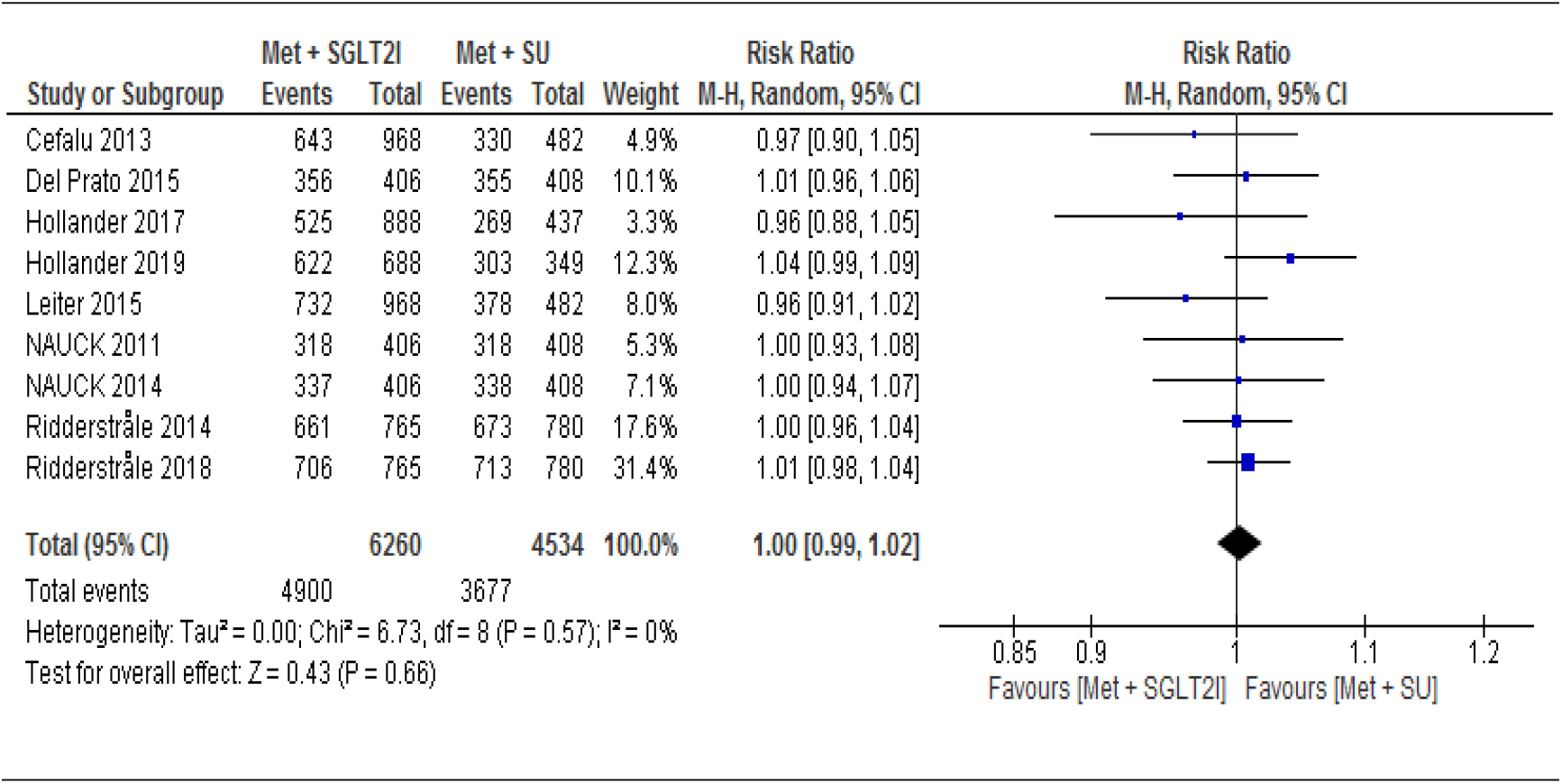
Comparison of the development of adverse events between patients who were on metformin-SGIT2I and metformin-sulphonylureas combination therapies. RR= Risk Ratio; CI= confidence interval; df= degrees of freedom; Met=Metformin; SGLT2I= Sodium-glucose co-transporter 2 inhibitor; SU= Sulfonylurea

#### Serious adverse events

All included trials assessed the risk of developing serious adverse events during the study period, where our analysis of the pooled data of these trials showed no statistically significant difference between the two groups (RR=0.96, 95% CI [0.79, 1.17], with statistically significant heterogeneity across trials (I^2^ = 68%, p=0.001) (Figure 7). Consistently, subgroup analysis showed no statistically significant difference between the two groups at year 1 (95% CI (RR=0.92 [0.53, 1.59], I^2^ = 75%), year 2 (RR=0.98 [0.69, 1.40], I^2^=80%), and year four (RR= 1.02 [0.87, 1.20], I^2^=0%), with subgroup differences of p = 0.92 and I^2^=0% (Appendix V).

**Figure 7:**
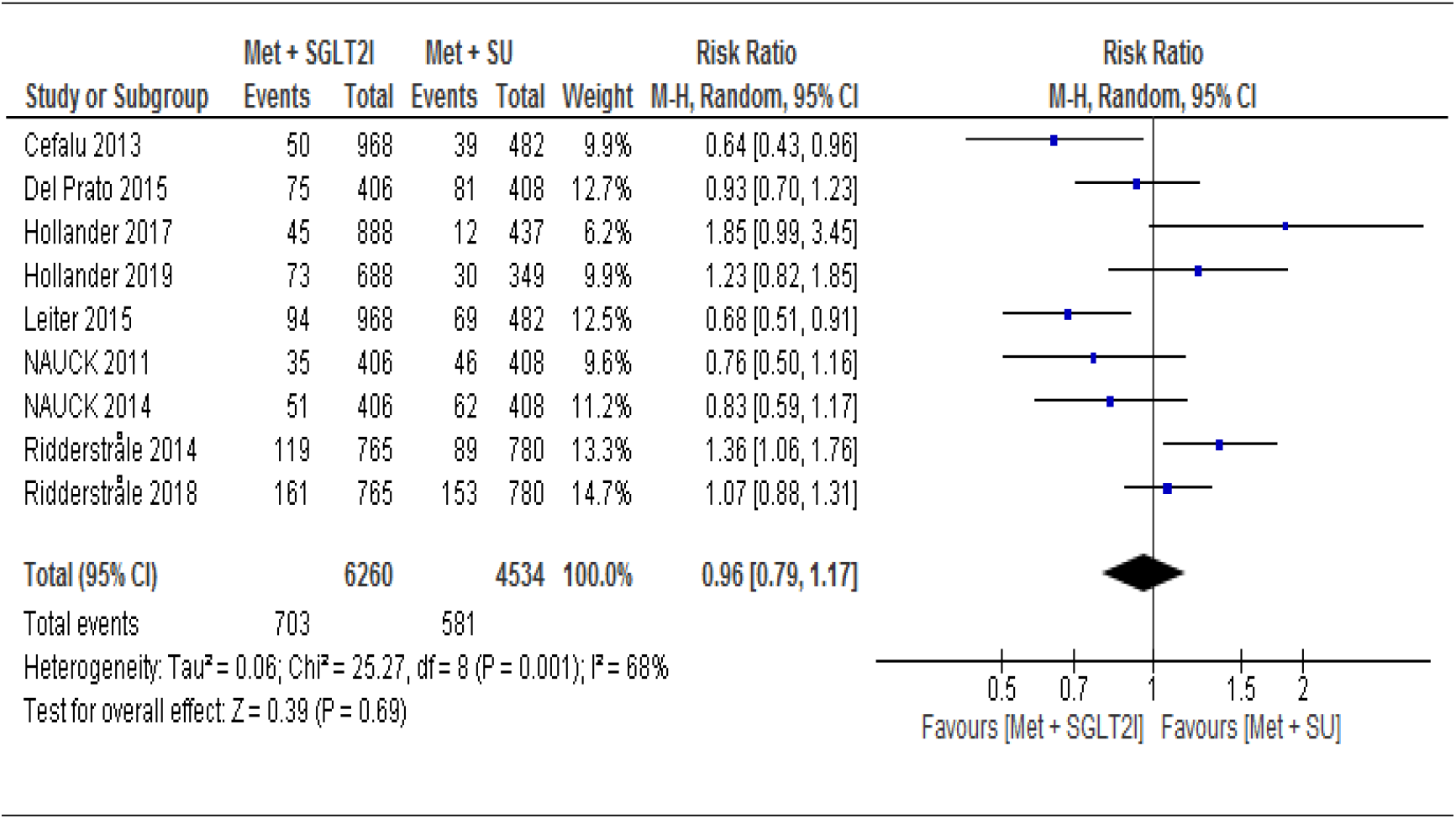
Comparison of the risk of developing serious adverse events between patients who were on metformin-SGIT2I and metformin-sulphonylureas combination therapies. RR= Risk Ratio; CI= confidence interval; df= degrees of freedom; Met=Metformin; SGLT2I= Sodium-glucose co-transporter 2 inhibitor; SU= Sulfonylurea

Five trials assessed the risk of serious adverse events related to study drug. The pooled result of these trials showed that serious adverse events related to study drug were less frequent on patients taking metformin-sulphonylureas combination, but the pooled result was not statistically significant at 95% CI (RR=1.34 [0.75, 2.36], I^2^= 0%) (Appendix VI).

#### Hypoglycemic events

We analyzed hypoglycemic events on pooled results of the nine trials involving 10,794 T2DM patients, considering the occurrence of at least one hypoglycemic event during the follow-up period. Patients under metformin-SGIT2I combination therapy were found to experience significantly fewer hypoglycemic events as compared to patients under metformin-sulphonylureas combination therapy (RR= 0.13, 95% CI [0.10, 0.17], P<0.00001) I^2^=67%, p=0.002) (Figure 8). Sensitivity analysis was performed by removing two low weighted trials [57,58]. However, the result of the remaining trials was similar to the nine trials for the risk of developing hypoglycemic events (RR= 0.13, 95% CI [0.10, 0.18, P<0.00001) with increased between study heterogeneity (I^2^= 73%, P=0009). To see the robustness of the result, we did subgroup analysis at different duration of follow-up. However, the risk of experiencing hypoglycemic events was consistently more frequent under patients on metformin-sulfonylureas combination therapy at 95% CI RR=0.12[0.08, 0.19], p<0.00001 at year one, RR=0.15[0.10, 0.22], p<0.00001 at year two and RR=0.10[0.07, 0.15], p<0.00001 at year four respectively. On the other hand, patients on metformin-SGLT2I were found to experience significantly higher genital mycotic infection than patients on metformin-sulfonylureas combination therapy RR=5.00, 95% CI [3.94, 6.33], p<0.00001 (Appendix VII).

**Figure 8:**
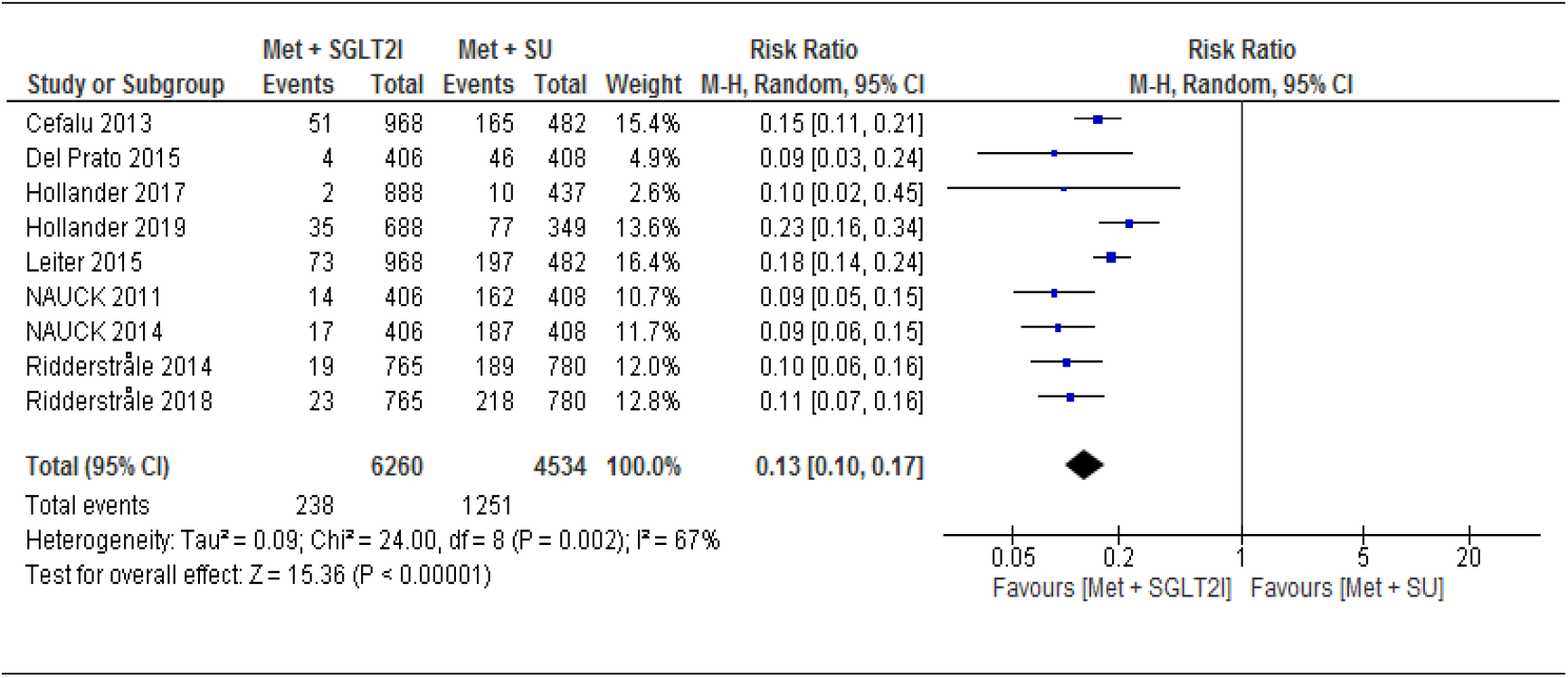
Comparison of the risk of developing hypoglycemic events between patients who were on metformin-SGIT2I and metformin-sulphonylureas combination therapies. RR= Risk Ratio; CI= confidence interval; df= degrees of freedom; Met=Metformin; SGLT2I= Sodium-glucose co-transporter 2 inhibitor; SU= Sulfonylurea

#### Glycosylated hemoglobin A1C

All the nine RCTs involving 9,180 participants determined the change in HbA1c (%) from baseline between the two arms. The pooled data of these trials showed significant difference in the mean difference of HbA1c between patients on metformin-SGLT2I and metformin-sulfonylureas combination MD= −0.10, 95% CI[−0.17, −0.03] %, p = 0.005), I^2^ = 63%, p=0.006) (Figure 9). Interestingly, subgroup analysis by duration of follow-up showed a reduction of HbA1c from baseline was not statistically significant between the two groups at year-one, with MD=-0.01[−0.13, 0.11] %, I^2^ = 67%. However, metformin-SGLT2I induced a greater reduction in HbA1c from baseline after 2 years (MD=-0.12[−0.20, −0.05] %, p=0.001) and after 4 years (MD=-0.24[−0.37, - 0.10] %, p=0.0007) (Appendix III).

**Figure 9:**
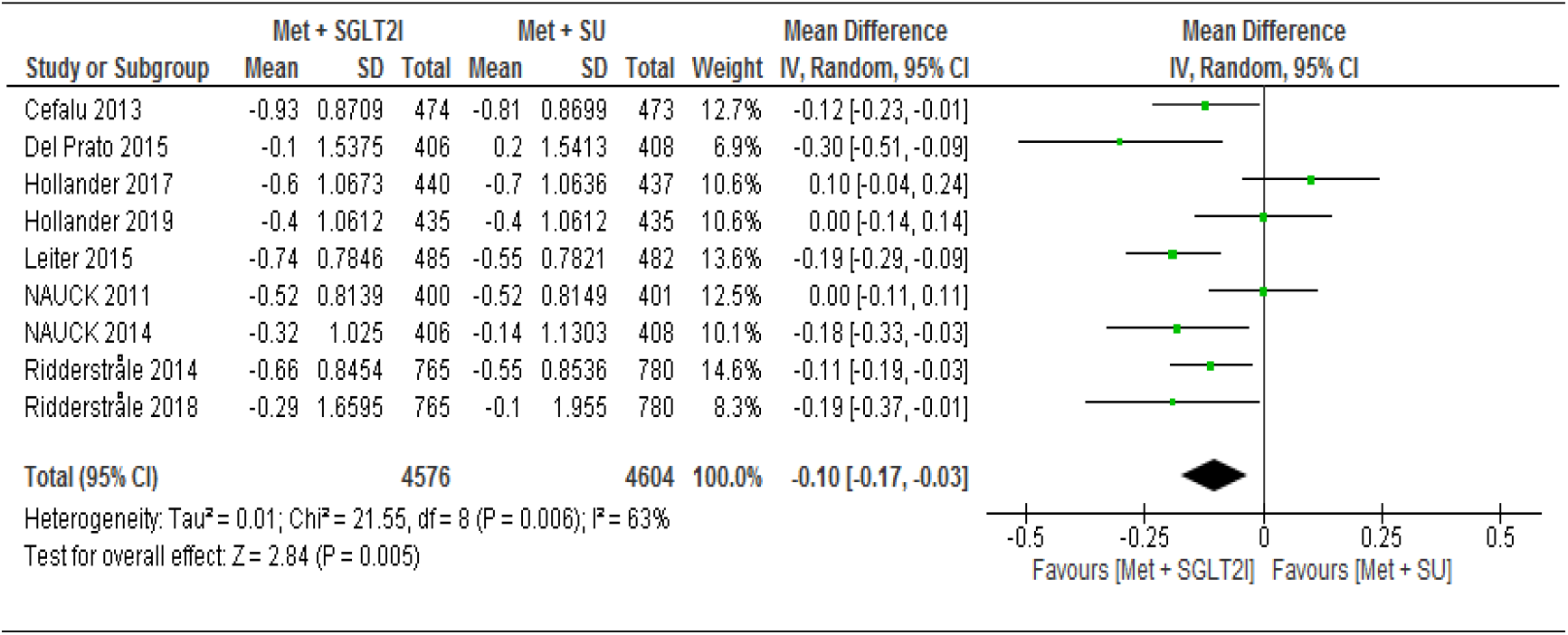
Comparison of change in HbA1c (%) from baseline between patients who were on metformin-SGIT2I and metformin-sulphonylureas combination therapies. SD= standard deviation; CI= confidence interval; MD= mean difference; Met=metformin; SGLT2I= Sodium glucose co-transporter 2 inhibitor; SU= sulfonylurea

#### Body weight

All trials included in meta-analysis assessed the change in body weight from baseline between the two groups. The pooled result showed that body weight of patients on metformin-SGLT2I was reduced by 4.57 kg from baseline compared to patients on metformin-sulfonylureas (MD= −4.57, 95% CI [−4.74, −4.39] kg, p < 0.00001), I^2^ = 0%) (Figure 10). We conducted a sensitivity analysis by removing the low weighted [55] and the high weighted [52] study and the result was consistent with the nine studies. Surprisingly, subgroup analysis also showed consistent results following different duration of follow-up that the mean difference of change in body weight from baseline at year one (MD=-4.52 [−4.79, −4.24] kg, p<0.00001, I^2^ = 0%); at year two (MD=-4.56 [−4.81, - 4.31] kg, p<0.00001, I^2^ = 0% and at year four (MD=-4.76[−5.27, −4.26] kg, p<0.00001, I^2^ = 0%) (Appendix IX).

**Figure 10:**
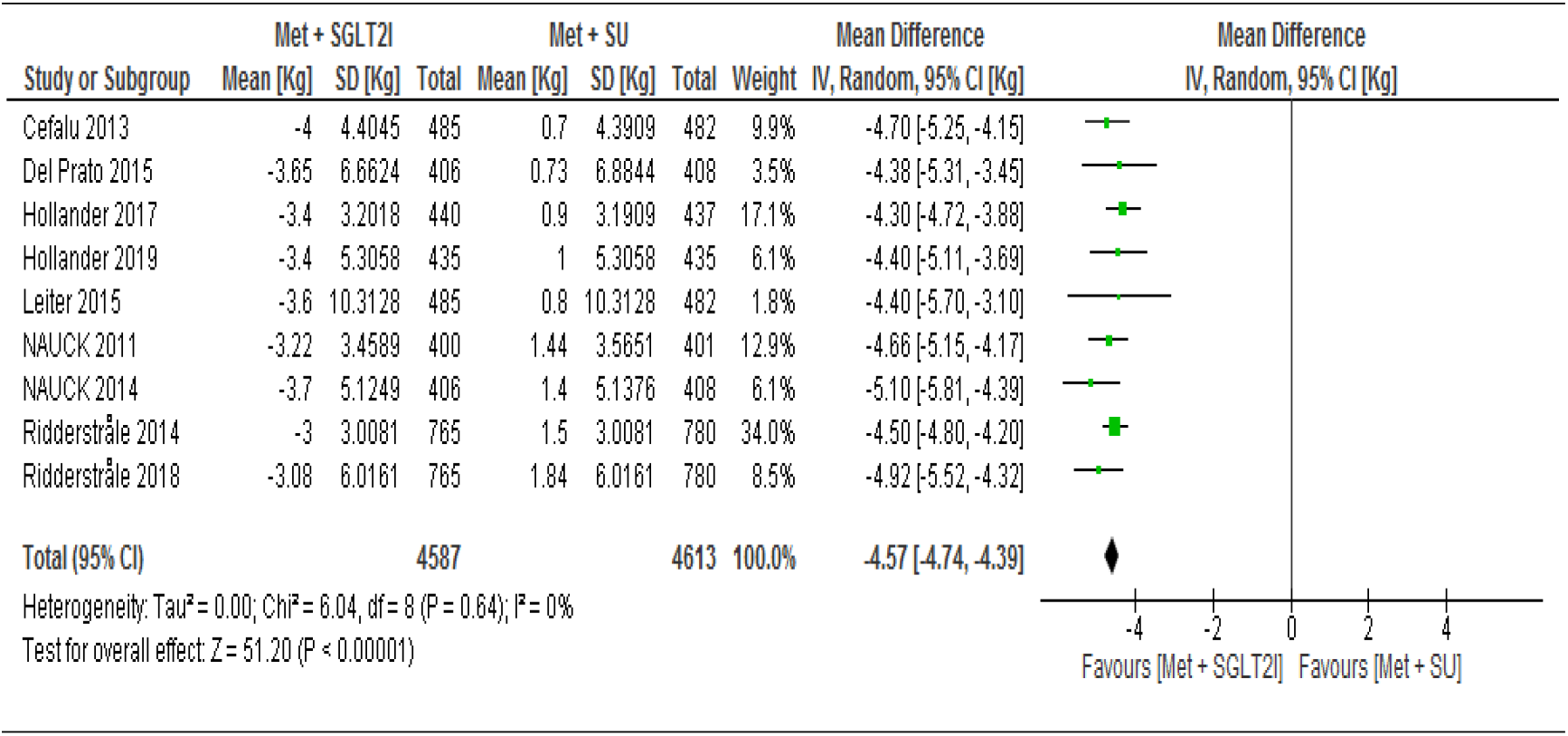
Comparison of change in body weight (Kg) from baseline between patients who were on metformin-SGIT2I and metformin-sulphonylureas combination therapies. SD= standard deviation; CI= confidence interval; MD= mean difference; Met= Metformin; SGLT2I= Sodium-glucose co-transporter 2 inhibitor; SU= Sulfonylurea

#### Fasting plasma glucose

Eight trials assessed the change in fasting plasma glucose level from baseline between the intervention and control. The pooled result showed that FPG level was reduced by 0.55 mmol/L from baseline with patients on metformin-SGLT2I combination therapies (MD= −0.55, 95% CI [−0.69, −0.41] mmol/L, p < 0.00001, I^2^ = 57%) (Figure 11). Sensitivity analysis was conducted by removing one outlier with a low weighted study [57]. But the result was consistent with the eight study (MD=-0.55, 95% CI [−0.67, −044], p < 0.00001). Subgroup analysis of seven trials also showed consistent results following different duration of intervention and control that the mean difference of change in FPG level from baseline at year one (MD=-0.47 [−0.63, −0.30] mmol/L, p<0.00001, I^2^ = 0%), at year two (MD=-0.56 [−0.74, −0.38] mmol/L, p<0.00001, I^2^ = 59% and at year four (MD=-0.70[−0.98, −0.42] mmol/L, p<0.00001) (Appendix X).

**Figure 11:**
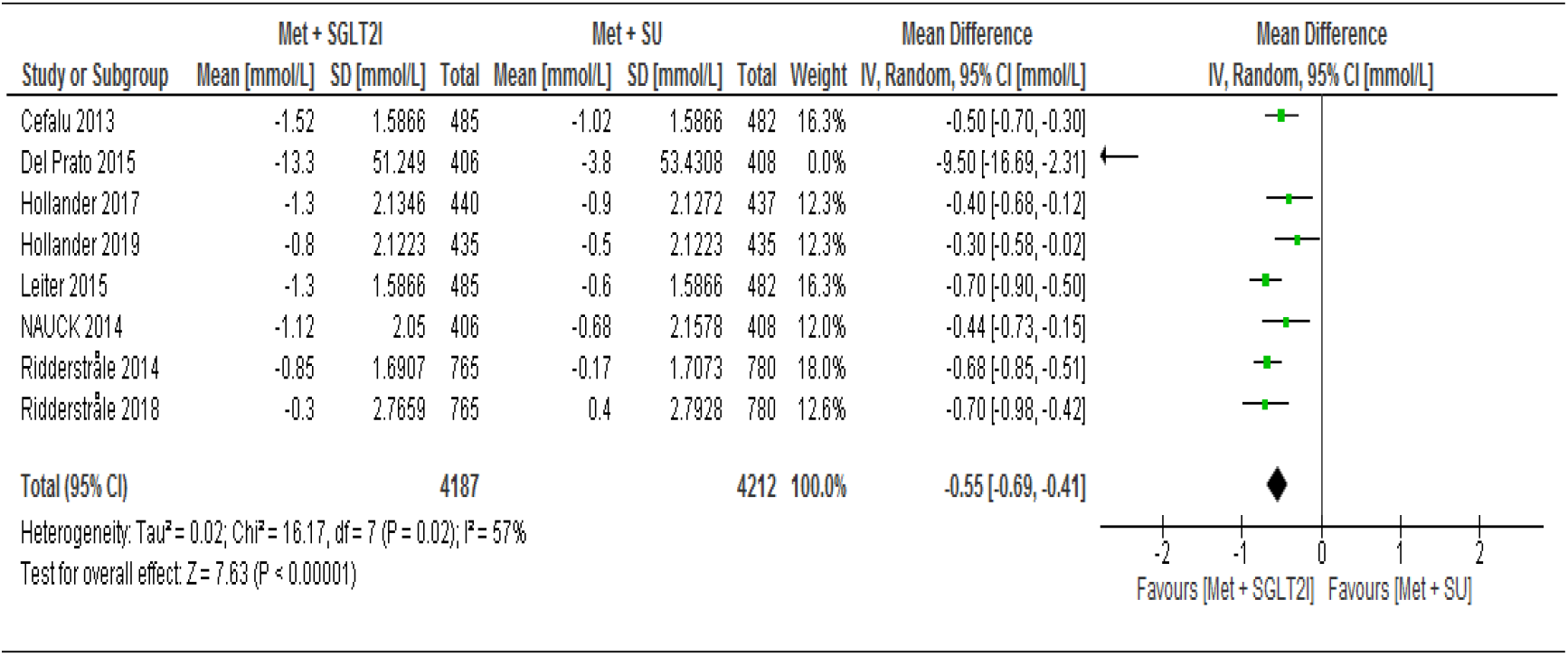
Comparison of change in FPG (mmol/L) from baseline between patients who were on metformin-SGIT2I and metformin-sulphonylureas combination therapies. SD= standard deviation; CI= confidence interval; MD= mean difference; Met= Metformin; SGLT2I= Sodium-glucose co-transporter 2 inhibitor; SU= Sulfonylurea

## Discussion

The result of this systematic review and meta-analysis showed that the risk of all-cause mortality/death events was not statistically significant between patients on metformin-SGLT2I and metformin-sulfonylureas combination therapy (RR = 0.93, 95% CI [0.52, 1.67], p = 0.81). In agreement with our findings a previous study reported non-significant difference in all-cause mortality and cardiovascular death between the two arms (RR=0.96, 95% CI [0.44, 2.09], P = 0.91[40]. Similarly, SCOUT trial [76] also revealed a lower and statistically nonsignificant cardiovascular death and all-cause mortality in T2DM patients who were on metformin-sulfonylurea combination therapy. However, another study [29] reported that the use of sulfonylureas as a second-line drug has a significantly associated with an increased risk of myocardial infarction and all-cause mortality.

The goal of durable glycemic control is to reduce the long-term risk of diabetes-related cardiovascular morbidity and mortality [56]. Recent studies showed that SGLT2Is have been shown to decrease cardiovascular events in treating high-risk patients with T2DM [34]. Likewise, the addition of empagliflozin to metformin therapy improves in patients with established CVD or heart failure [77]. Even though the current meta-analysis of metformin-SGLT2I combination showed a favorable effect on cardiovascular outcomes (acute myocardial infarction, aortic aneurism, coronary artery occlusion, and atherosclerosis), the pooled analysis was not statistically significant.

Lowering blood pressure is significantly important to reduce the risk of CVD and death in many patients with T2DM. A 10 mmHg reduction in systolic blood pressure decreased the risk of major CVD events by 20% [78]. SGLT2I induced a greater reduction in systolic and diastolic blood pressure [72]. Similar with the other findings, the pooled result of our study showed a reduction in both systolic and diastolic blood pressure by 4.77 mmHg and 2.07 mmHg, respectively, with patients on metformin-SGLT2I combination therapy as compared with metformin-sulfonylureas, which might be due to the effect of SGLT2I reducing renal glucose reabsorption and increasing urinary excretion [50]. In consistent with our findings, previous studies reported a greater increases from baseline in HDL-C and LDL-C in patients who were on SGLT2I groups as compared with sulfonylureas [50, 53, 55].

The result of this meta-analysis showed that the overall incidence of AEs was similar across the two arms, which is in consistent with a trial reported by elsewhere [58]. However, metformin-SGLT2I combination was associated with a higher episode of GMI which is similar to other meta-analysis reported an increased risk of GMI with SGLT2Is [72]. In addition, for both genders, the proportion of patients reporting symptoms of GMI was higher in the SGLT2I group than in the sulfonylureas [57]. However, these infections respond to standard antimicrobial treatment and their incidence declines with time [56]. SAEs such as ketoacidosis, bone fractures, and pyelonephritis were rarely reported with metformin-SGLT2I [52], while the pooled result of the current study did not find a statistically significant difference between metformin-SGLT2I and metformin-sulfonylureas combinations. Even though the pooled analysis did not find statistically significant results, AEs related to study drug was observed more in patients with metformin-SGLT2I, but more SAEs related to study drug in metformin-sulfonylureas. This might be due to the hypoglycemic effect of sulfonylureas and GMI as a result of SGLT2I respectively.

Sulfonylureas as second-line drugs are associated with an increased risk of severe hypoglycemia [78], and our study is in support of this. The hypoglycemic effect of sulfonylureas might be due to its insulin-dependent mechanism of action, while less hypoglycemic effect of SGLT2Is is due to the non-insulin dependent mechanism of action.

Obesity is one of the main risk factors for T2DM and representing a major worldwide health problem. Lowering body weight is an important part of T2DM management. SGLT2I has been associated with an added benefit of weight loss in patients with T2DM, whereas sulfonylureas are reported to increase body weight [79]. In support of this evidence, our study showed a 4.57 kg weight loss in metformin-SGLT2I arm than the metformin-sulfonylureas. The weight loss caused by SGLT2I is probably due to the loss of calories via urine and glucose-induced osmotic diuresis [50].

Long term glycemic control is the major goal of diabetes management to prevent both microvascular and macrovascular complications of diabetes mellitus [80]. Both metformin-SGLT2I and metformin-sulfonylureas combinations are effective to control HbA1c for a short duration of follow-up. However, for a long duration of follow-up, metformin-SGLT2I are more effective [72]. Our finding is consistent with this evidence where both groups were equally effective for a one year duration of follow-up; however, as the duration of follow-up increases to 4 years, metformin-SGLT2I combination showed a significant reduction of HbA1c. Eight previously conducted trials reported a higher reduction of FPG in patients randomized to metformin-SGLT2I [50, 51-53, 55-58], and our pooled result is in support of their finding as it showed a 0.55 mmol/L reduction of FPG under patients on metformin-SGLT2I.

This study reported important information about the cardiovascular safety and efficacy between the two combination therapies. However, we acknowledge that available studies are very limited and heterogeneous. There remains a need for additional long-term trials comparing the overall safety, efficacy, and cost-effectiveness of metformin-SGLT2Is and metformin-sulfonylureas combination therapies.

## Conclusion

A combination therapy of metformin and sodium-glucose cotransporter-2 inhibitors is a safe and efficacious alternative to combination therapy of metformin and sulphonylureas for patients with T2DM who are at risk of cardiovascular comorbidity. However, there remains a need for additional long-term randomized controlled trials as available studies are very limited and heterogeneous.

## Data Availability

All relevant data are within the manuscript and its supporting information files.

## Abbreviations

AEs: Adverse Events
AMI: Acute Myocardial Infraction
CAD: Coronary Artery Disease
CAO: Coronary Artery Occlusion
CVD: Cardiovascular Disease
DBP: Diastolic Blood Pressure
FPG: Fasting Plasma Glucose
GMI: Genital Mycotic Infection
HbA1C: Hemoglobin A1C
HDL-C: High Density Lipoprotein Cholesterol
LDL-C: Low Density Lipoprotein Cholesterol
MD: Mean Difference
MI: Myocardial Infraction
RCT: Randomized Controlled Trials
RR: Risk Ratio
SAEs: Serious Adverse Events
SBP: Systolic Blood Pressure
SGLT2I: Sodium Glucose Co-transporter 2 Inhibitor
T2DM: Type 2 Diabetes Mellitus

## Declarations

### Ethics approval and consent to participate

Not applicable.

### Consent for publication

Not applicable.

### Availability of data and materials

All relevant data are within the manuscript and its supporting information files.

### Competing interests

All review authors declare that they have no competing interests.

### Funding

None.

### Authors’ contributions

DG (first author) was responsible for the conception of review protocol, study design, literature search, data extraction, quality assessment, data analysis, interpretation, and drafting the manuscript. DG (second author) and TM were responsible for the quality assessment and reviewing the manuscript. All authors have read and approved the manuscript.

## Acknowledgement

Not applicable.

## Notes

### Competing Interest Statement

The authors have declared no competing interest.

